# Mapping disparities in viral infection rates using highly-multiplexed serology

**DOI:** 10.1101/2024.02.22.24303200

**Authors:** Alejandra Piña, Evan A Elko, Rachel Caballero, Mary Mulrow, Dan Quan, Lora Nordstrom, John A Altin, Jason T Ladner

## Abstract

Despite advancements in medical interventions, the disease burden caused by viral pathogens remains large and highly diverse. This burden includes the wide range of signs and symptoms associated with active viral replication as well as a variety of clinical sequelae of infection. Moreover, there is growing evidence supporting the existence of sex– and ethnicity-based health disparities linked to viral infections and their associated diseases. Despite several well-documented disparities in viral infection rates, our current understanding of virus-associated health disparities remains incomplete. This knowledge gap can be attributed, in part, to limitations of the most commonly used viral detection methodologies, which lack the breadth needed to characterize exposures across the entire virome. Additionally, virus-related health disparities are dynamic and often differ considerably through space and time. In this study, we utilize PepSeq, an approach for highly-multiplexed serology, to broadly assess an individual’s history of viral exposures, and we demonstrate the effectiveness of this approach for detecting infection disparities through a pilot study of 400 adults aged 30-60 in Phoenix, AZ. Using a human virome PepSeq library, we observed expected seroprevalence rates for several common viruses and detected both expected and previously undocumented differences in inferred rates of infection between our Hispanic White and non-Hispanic White individuals.

**Importance:** Our understanding of population-level virus infection rates and associated health disparities is incomplete. In part, this is because of the high diversity of human-infecting viruses and the limited breadth and sensitivity of traditional approaches for detecting infection events. Here, we demonstrate the potential for modern, highly-multiplexed antibody detection methods to greatly increase our understanding of disparities in rates of infection across subpopulations (e.g., different sexes or ethnic groups). The use of antibodies as biomarkers allows us to detect evidence of past infections over an extended period of time, and our approach for highly-multiplexed serology (PepSeq) allows us to measure antibody responses against 100s of viruses in an efficient and cost-effective manner.

## Introduction

Despite advancements in the development of medical interventions, the global burden of disease caused by viral pathogens remains substantial and highly diverse. This burden includes a wide range of morbidities associated with active viral replication ranging in severity from fever, muscle aches, and rash to encephalitis, immunosuppression, respiratory failure, and congenital birth defects (1–4). Additionally, it includes an array of clinical sequelae of infection (e.g., Guillain-Barre syndrome, Multisystem inflammatory syndrome, Long COVID), many of which remain poorly understood (5–7). Viral infections have even been linked to the onset of a number of non-communicable diseases, such as myocarditis (8), diabetes (9), celiac disease (10), obesity (11), multiple sclerosis (12, 13), cancer (14), and Alzheimer’s disease (15).

Although the health effects caused by viral infections are observed widely in the general population, currently documented national trends highlight several sex– and ethnicity-based health disparities in the prevalence of viral infections and thus, virus-associated disease. For example, in the United States (US), seroprevalence of human papillomavirus (HPV) and herpes simplex virus 2 (HSV-2) have been shown to be roughly twice as high among women compared to men (16, 17). In contrast, human immunodeficiency virus 1 (HIV-1) disproportionately affects men in the US, particularly men who have sex with men. In 2021, 69% of new HIV-1 diagnoses in the US were among men who have sex with men, despite this group representing only 3.9% of the US population (18, 19). Ethnic disparities have also been documented for HIV-1 in the US. A 2015-2019 CDC surveillance report showed the incidence of HIV-1 infection among Hispanics was four times higher than that among non-Hispanic Whites. Furthermore, disparities in viral infections can vary in space and time. For example, over a 12 year period the average annual incidence per 100,000 people of hepatitis A virus (HAV) in a Native American population in Arizona decreased from 289 to 6 due to vaccination efforts (20). Vector-borne viruses provide striking examples of geographical disparities. For example, dengue virus is transmitted by mosquitoes of the genus *Aedes* that live primarily in tropical regions (21). In 2020 the number of locally transmitted cases of dengue virus was 0.025 per 100,000 people in the continental United States, compared to 23.68 cases per 100,000 people in Puerto Rico (22).

Despite these well documented disparities in viral infection rates, our understanding of virus-associated health disparities remains incomplete. In part, this is because the most commonly used methodologies for detecting viral infections are limited in their breadth, both in terms of the number of viruses they can detect and/or the period of time during which detection is possible. Molecular assays detect viral nucleic acids and therefore lack sensitivity in cases where the infection has already been cleared or if the sampled fluid does not contain the virus (23). In contrast, serological assays detect antiviral antibodies that can persist for years after exposure due to the body’s long-lived humoral immune response. However, the most commonly used serological assays, such as the Enzyme-Linked Immunosorbent Assay (ELISA), only test for one virus (and typically one protein) at a time (24).

Recent advances in serological methods have overcome previous limitations in breadth and are enabling unprecedented views into the viral exposure histories of individuals (25–28). Using current approaches for highly-multiplexed serology (e.g., PepSeq (29), PhIP-Seq (30)) it is now possible to characterize antibody binding to 100,000s of antigens in a single assay using <1 µL of blood. In this study, we utilize the PepSeq platform to demonstrate the potential of highly-multiplexed serology to broadly characterize differences in seroprevalence across the human virome between different demographic groups. Through the characterization of antiviral antibodies in samples collected over ∼1 month within a single healthcare system in Phoenix, AZ, we document significant differences in infection rates between the Hispanic White (HW) and non-Hispanic White (NHW) populations for several viruses, including some that are rarely included in population-level surveys.

## Results

### Study population

Between late May and early June 2020, we collected 400 remnant serum samples from 11 Valleywise Health facilities, a large safety net hospital system in Phoenix, AZ. These samples were distributed equally among four subpopulations: 100 HW men, 100 HW women, 100 NHW men and 100 NHW women. To minimize the impact of age-related differences in seropositivity, we limited our focal age range to 30-60 years, and the age distributions were not significantly different (t-test) between genders or ethnicities, with mean age ranging between 45.1 and 46.2 (Figure 1A). We also investigated payor source for each sampled individual (Figure 1B), as this can serve as a proxy for socioeconomic status (31). Overall, the vast majority of the individuals included in our study were either covered by a government insurance plan (52.8%; Tricare, Medicare and/or Medicaid) or were uninsured (31.3%; self-pay). Only ∼13.1% of the individuals in our study were covered by commercial insurance plans and this percentage did not vary considerably among our subpopulations, though we observed a slightly higher rate of commercial payor for NHW females (20.2%) compared to the other three groups (10-11.2%). However, we did observe substantial differences among our subpopulations in the proportions covered by either government programs or uninsured. The NHW populations were covered in higher proportions by government insurance programs (HW-M: 44.9%, HW-F: 16%, NHW-M: 79.8%, NHW-F: 72.7%), while the HW populations were more likely to be uninsured (HW-M: 39.8%, HW-F: 72%, NHW-M: 10.1%, NHW-F: 3%).

**Figure 1.**
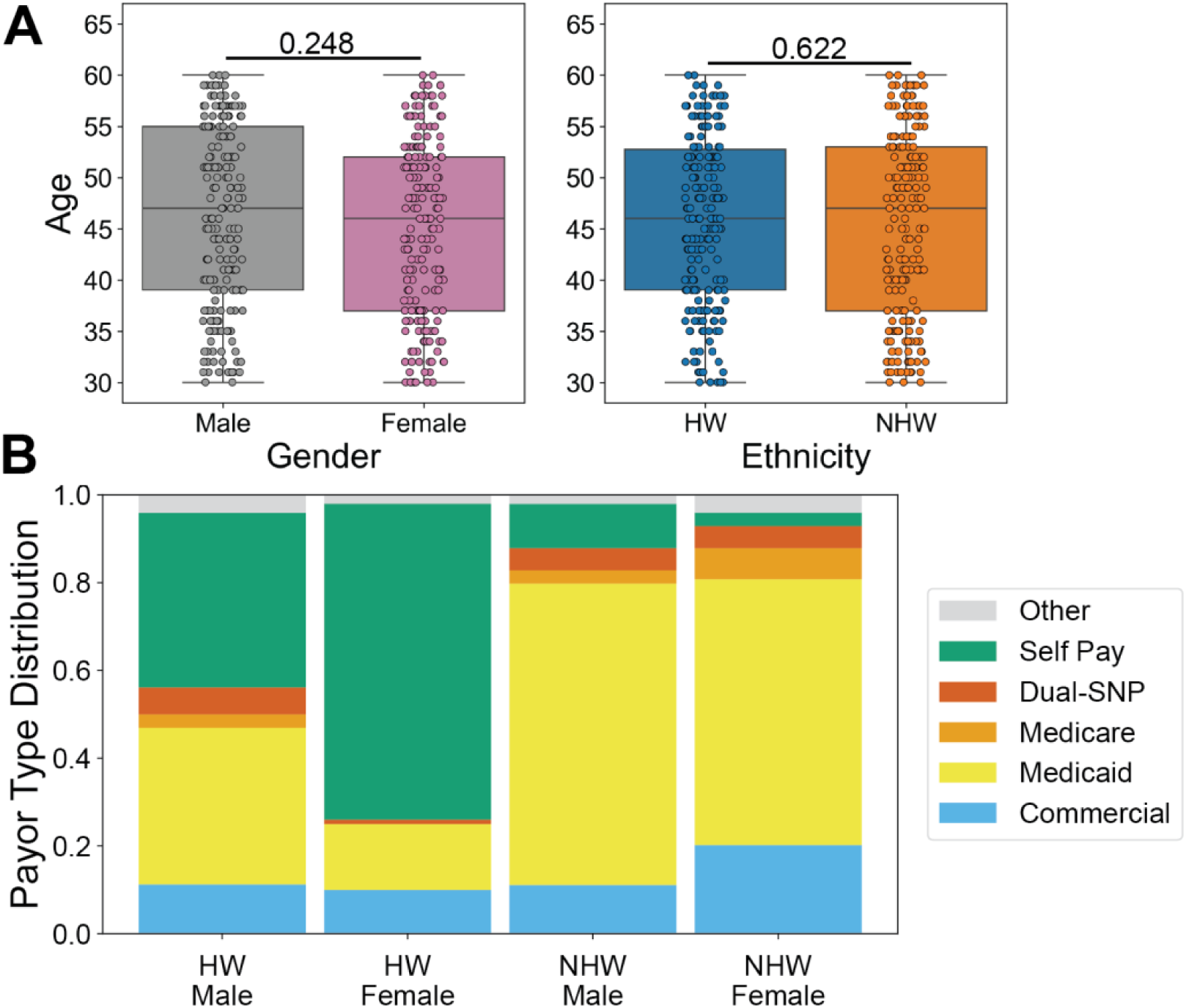
Demographics of the study population. Remnant serum samples were collected from 400 individuals at Valleywise Health in Phoenix, AZ and assayed using the HV1 PepSeq library. Four samples were excluded from the analysis due to poor correlation between replicates. A) The study population ranged in age from 30-60 years old and there was no statistically significant difference in the age distributions between genders and ethnicities (t-tests). Individual t-test p-values comparing Male/Female and HW/NHW ages are indicated above the respective plots. Each circle represents an individual. The line within each box represents the median, while the lower and upper bounds of each box represent the 1st and 3rd quartiles, respectively. The whiskers extend to points that lie within 1.5 interquartile ranges of the 1st and 3rd quartiles. B) Payor source varied substantially among focal subpopulations. Payor source was consolidated into six general categories (Table S5) and is shown for each subpopulation. Dual-SNP refers to any dual special needs plans for individuals who qualify for both Medicare and Medicaid.

### PepSeq Analysis

All 400 serum samples were assayed, in duplicate, using our human virome version 1 (HV1) PepSeq library (26), and we obtained an average of 2.2M Illumina sequencing reads per sample, which equates to an average of 9.2 reads per unique HV1 peptide per sample. Four samples were excluded from further analysis due to lower than normal correlation between replicates, which may indicate the occurrence of molecular bottlenecks or contamination of one of the replicates while performing the assay (29). These included two HW males, one NHW male and one NHW female. Therefore, all the analyses presented here include a total sample size of 396.

Based on visual comparisons of experimental samples and buffer-only negative controls (Supplemental Figure 1), as well the analysis of a separate group of negative controls that were not considered in the formation of bins or normalization of the data, we chose a set of four different Z score thresholds (10, 15, 20 and 25) for identifying enriched peptides. Higher thresholds are expected to have reduced sensitivity, but increased specificity. To estimate the false positive rate at each of these thresholds, we analyzed all pairwise combinations of 9 buffer-only negative controls (n=36). We observed an average of 5.5, 1.1, 0.28 and 0.17 putatively enriched peptides from these control pseudoreplicate analyses for thresholds of 10, 15, 20 and 25, respectively. In contrast, from the assays of serum samples, we observed an average of 1335, 1071, 926 and 828 enriched peptides for thresholds of 10, 15, 20 and 25, respectively. This equates to expected false positive rates of approximately 0.41%, 0.1%, 0.03% and 0.02%, respectively.

To broadly characterize enrichment patterns within our data set, we averaged the number of enriched peptides across all Z score thresholds for each sample. We did not observe significant differences between the average number of enriched peptides by gender (F=1009.92, M=1070.30; t-test p-value=0.135) or ethnicity (HW=1044.37, NHW=1035.55; t-test p-value=0.827) (Figure 2A). We also observed no significant correlation between the number of enriched peptides and age (Figure 2B; Pearson correlation p-value=0.221).

**Figure 2.**
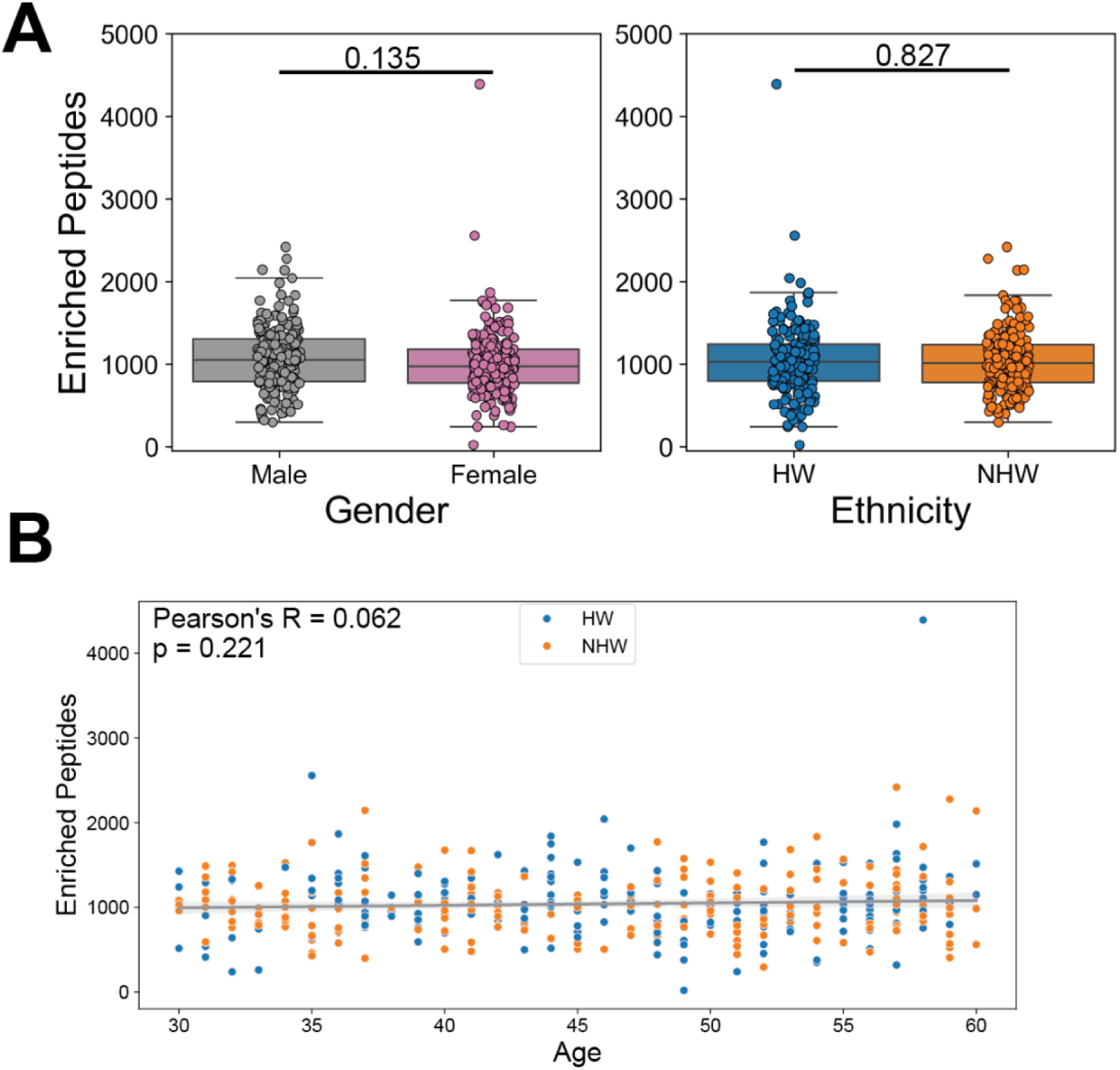
PepSeq identifies similar overall levels of antibody reactivity against viral peptides between genders and ethnicities. A) Box plots depicting the average number of enriched PepSeq peptides for each sample across the four Z score thresholds (Z = 10, 15, 20, and 25). Average number of enriched peptides: Female=1009.92, Male=1070.30, HW=1044.37, NHW=1035.55. Individual t-test p-values comparing Male/Female and HW/NHW enriched peptide counts are indicated above the respective plots. Each circle represents an individual. The line within each box represents the median, while the lower and upper bounds of each box represent the 1st and 3rd quartiles, respectively. The whiskers extend to points that lie within 1.5 interquartile ranges of the 1st and 3rd quartiles. B) Scatter plot with best-fit line showing average number of enriched peptides by age. Ethnicity is indicated by the color of the points, HW=blue and NHW=orange. Gray diagonal line indicates the best-fit linear regression with the shaded gray areas showing the 95% confidence interval. Pearson correlation was used to test for significance (p-value=0.221).

The enriched peptides for each Z score threshold were then converted into putative virus species-level serostatus calls using two different sets of potential viruses: one included all 390 HV1 target species as viruses to which the study individuals may have been exposed (“Full”), while the other only included 93 viruses (“Focal93”), excluding those that are unlikely to have been encountered by our focal population (Table S1). Overall, in comparing the results from these two different sets of potential viruses, we observed strong positive correlations in individual-level estimates of the number of seropositive species (Pearson’s correlation coefficients: 0.957 – 0.963; p-values: 1.13e^-214^ – 2.50e^-226^) and in species-level estimates of seroprevalence (Pearson’s correlation coefficients: 0.983 – 0.997; p-values: 7.35e^-60^ – 2.69e^-87^). As expected, we also found that most of the seropositive calls using the Full species set were for viruses included in our Focal93 list (76%, 82%, 86%, and 89% for Z score thresholds of 10, 15, 20, and 25, respectively), despite the fact that the Focal93 viruses make up a minority of the Full list of virus species (24%). The relatively small number of predicted exposures to viruses outside of our Focal93 subset are likely due to the presence of cross-reactive antibodies and/or true exposure to unusual or uncharacterized viruses. Separately for each set of viruses, we averaged the number of species per sample across all Z score thresholds to broadly characterize patterns within our subpopulations, and we found no significant differences (t-test) in the average number of predicted seropositive species by gender (Focal93: F=29.44, M= 29.92; Full: F=33.58, M=34.11) or by ethnicity (Focal93: HW=29.84, NHW=29.52; Full: HW=34.20, NHW=33.49) (Figure 3A). However, we did observe a significant positive correlation between age and the number of predicted seropositive species from both sets of potential viruses (Focal93: Pearson correlation p-value=0.007, Figure 3B; Full: Pearson correlation p-value=0.001, not shown).

**Figure 3.**
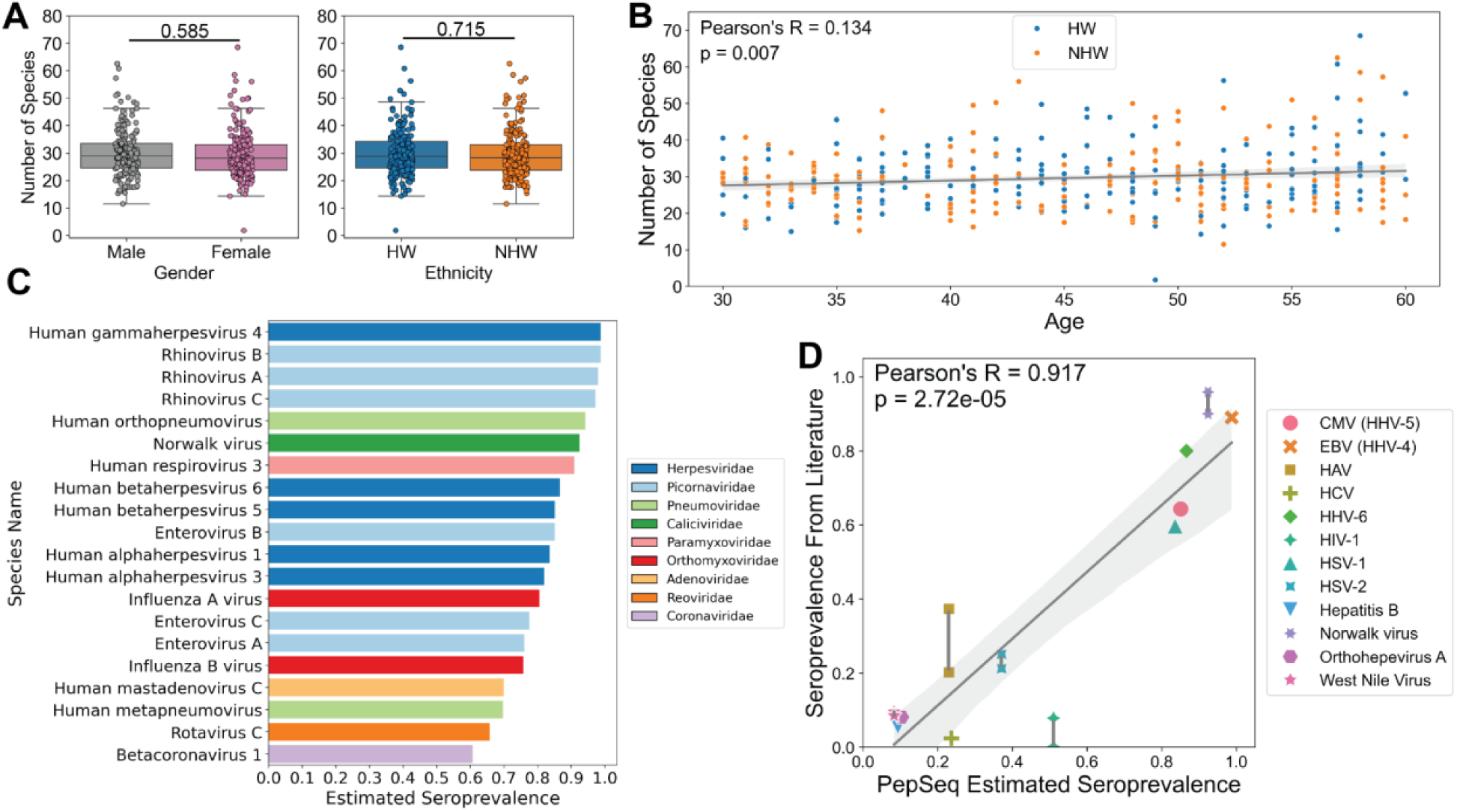
PepSeq-based estimates of seropositivity correlate with age and independent estimates from singleplex assays. A) Box plots depicting the number of virus species selected as seropositive by the PepSIRF *deconv* algorithm for each sample (Focal93 linkage map), divided according to gender (left) and ethnicity (right). Average number of putatively seropositive virus species: Female=29.44, Male=29.92, HW=29.84, NHW=29.52. T-tests comparing Male/Female and HW/NHW were non-significant (p-values indicated above the respective plots). Each circle represents an individual. The line within each box represents the median, while the lower and upper bounds of each box represent the 1st and 3rd quartiles, respectively. The whiskers extend to points that lie within 1.5 interquartile ranges of the 1st and 3rd quartiles. B) Scatter plot with best-fit line showing average number of predicted seropositive virus species by age. Ethnicity is indicated by the color of the points, HW=blue and NHW=orange. Pearson correlation was used to test for significance (p-value = 0.007). Gray diagonal line indicates the best-fit linear regression with the shaded gray areas showing the 95% confidence interval. C) Estimated seroprevalence for the 20 most prevalent virus species across the entire patient cohort. Bar colors indicate the viral family. D) PepSeq estimated seroprevalence across the full dataset compared to published seroprevalence values from studies in the United States (Table S8). Different estimates for the same virus are connected by a vertical line. The gray diagonal line indicates the best-fit linear regression with the shaded gray areas showing the 95% confidence interval. Pearson’s R value and p-value shown in the top left. Abbreviations: RSV=respiratory syncytial virus, EBV=Epstein-Barr virus, HHV=Human herpesvirus HSV=herpes simplex virus, CMV=cytomegalovirus, HAV=hepatitis A virus, HIV=human immunodeficiency virus, HCV=hepatitis C virus.

Next, to assess the performance of PepSeq in determining population level seroprevalence across a wide range of virus species, we calculated the estimated seroprevalence for the full cohort of 396 samples. This analysis resulted in the estimated seroprevalence for 306 virus species. Among the 20 virus species with the highest seroprevalence, the *Herpesviridae* and *Picornaviridae* families were the most frequently represented (Figure 3C). Seven virus species had an estimated seroprevalence greater than 0.9 (90%; Human gammaherpesvirus 4, Rhinovirus A-C, Human orthopneumovirus, Norwalk virus and Human respirovirus 3) (Figure 3C). For 12 virus species with published seroprevalence studies in the United States, we then compared the PepSeq estimates of seroprevalence with the published estimates based on more traditional singleplex assays (32–41). There was a highly significant correlation between the PepSeq estimated seroprevalence and the estimated seroprevalence found in the literature (p-value=2.72e-5, Pearson R=0.917) (Figure 3D).

### Identification of disparities

To identify statistically significant differences in estimated seroprevalence among our subpopulations, we fit a binomial generalized linear model (GLM) with a single dependent variable (seroprevalence) and three independent variables (ethnicity, gender, and age). The results of this analysis using the Full and Focal93 virus sets were remarkably consistent, and all the viruses that exhibited significant differences between subpopulations were present within the Focal93 subset (Table S2). Therefore, we only present the results for the Focal93 set of viruses.

In general, across virus species, we observed higher total seropositivity in older individuals, but there was no consistent directional change associated with gender or ethnicity (Supplemental Figure 2). However, no individual viruses exhibited significant correlations between age and seropositivity after correcting for multiple tests, which may be related to the limited range of ages included in this study (30-60 years old). We also found no significant differences in estimated seroprevalence between genders. However, we did observe several viruses with significant correlations between seropositivity and ethnicity, and these patterns were generally consistent across our four Z score thresholds (Figure 4A).

**Figure 4.**
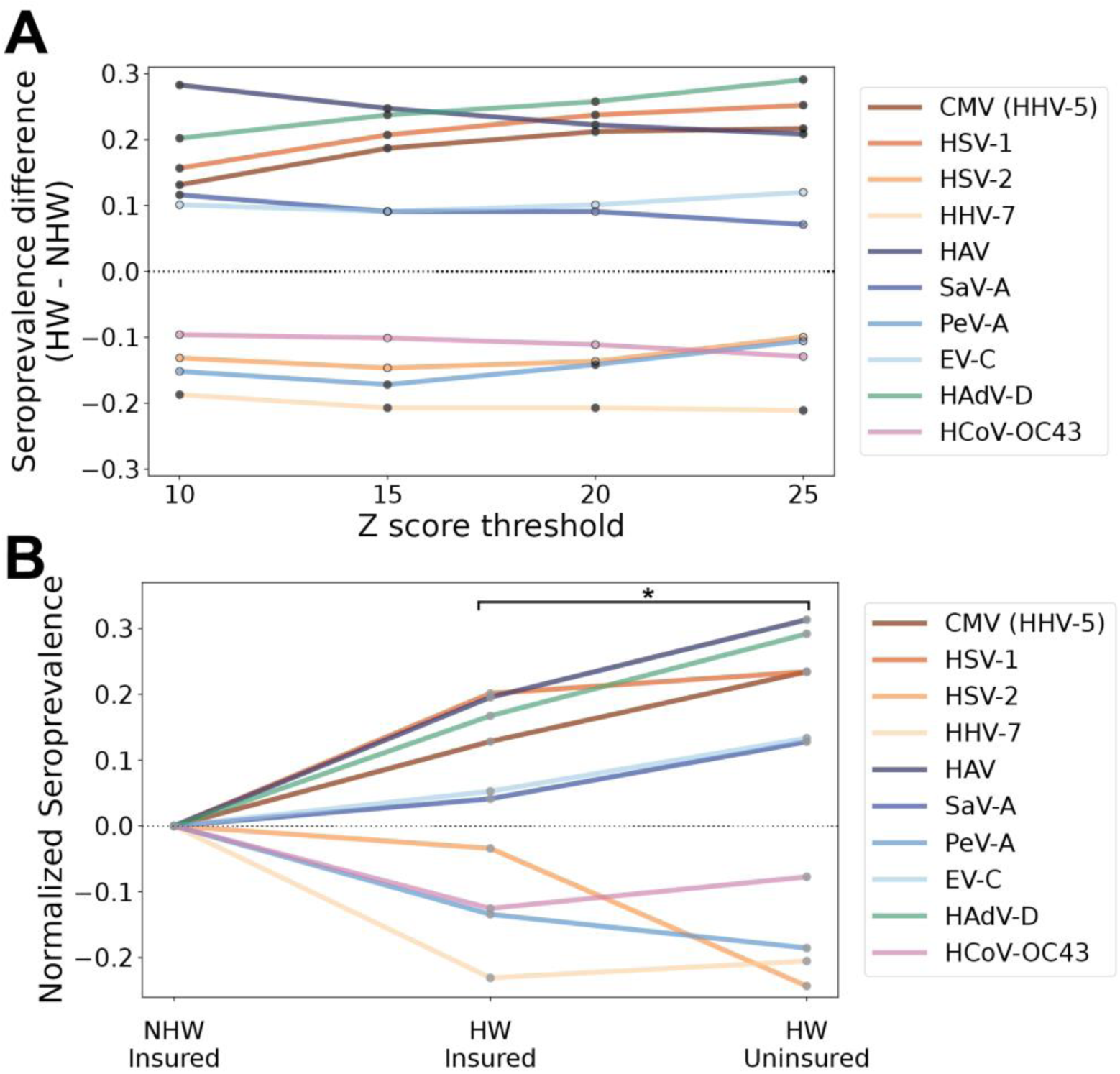
Significant differences in seroprevalence by ethnicity and payor status. A) Line plot depicting ten species with a significant difference (p-value<0.05) in seropositivity between HW and NHW calculated by fitting a generalized linear model at each Z score threshold before (outlined points) and after (filled points) Bonferroni correction for multiple tests. Negative differences indicate higher seroprevalence in NHWs and positive differences indicate higher seroprevalence in HWs. B) Line plot depicting normalized seroprevalence for the same ten viruses shown in (A), with values calculated separately for insured and uninsured individuals. Seroprevalence is being shown for a Z score threshold of 15 and was normalized against the value for insured NHWs. Asterisk indicates the significant increase of the absolute value of the normalized seroprevalence for uninsured HWs compared to insured HWs across all 10 viruses (paired t-test p-value=0.013). Abbreviations: CMV=cytomegalovirus, HSV=herpes simplex virus, HHV=human herpesvirus, HAV=hepatitis A virus, SaV-A = salivirus A, PeV-A=parechovirus A, EV-C=*Enterovirus C*, HAdV-D=human adenovirus D, HCoV-OC43=human coronavirus OC43.

In total, ten virus species exhibited ethnicity p-values<0.05 across all four Z score thresholds: cytomegalovirus (CMV, aka human herpesvirus 5), herpes simplex virus 1 (HSV-1) and 2 (HSV-2), human herpesvirus 7 (HHV-7), hepatitis A virus (HAV), salivirus A (SaV-A), parechovirus A (PeV-A), *Enterovirus C* (EV-C), human adenovirus D (HAdV-D) and human coronavirus OC43 (HCoV-OC43). After Bonferroni correction for multiple tests, seven of these remained significant at ≥1 Z score threshold and five remained significant across all four Z score thresholds (Figure 4A). Within the HW subpopulation, we observed significantly higher seropositivity for CMV, HSV-1, HAdV-D and HAV across all four Z thresholds and for SaV-A at a Z threshold of 10 (Figure 4A). Within the NHW subpopulation, we observed significantly higher seropositivity for HHV-7 across all thresholds and for PeV-A at Z thresholds of 15 and 20.

For all ten viruses showing significant or nearly significant differences in seroprevalence between ethnicities, we also looked at the relationship between seropositivity and insurance status. Specifically, using a Z score threshold of 15, we compared seroprevalence estimates among three subsets of our study population: insured NHWs (n=185), insured HWs (n=87), and uninsured HWs (n=111). Uninsured NHWs were excluded because of a low sample size (n=13), but generally showed seroprevalence estimates similar to insured NHWs. Our results showed that for most viruses, estimated seroprevalence within insured HWs was intermediate between the NHW insured and HW uninsured subpopulations (Figure 4B). In fact, when considering all ten viruses together, we observed a significant difference between the normalized estimated seroprevalences in the insured HW group compared to the uninsured HW group (absolute values, paired t-test p-value=0.013). For all six of the viruses with higher seropositivity among HWs, seropositivity was highest among uninsured individuals, and for half (2/4) of the viruses with lower seropositivity in HWs, seropositivity was lowest in the HW uninsured group. Two viruses, HHV-7 and HCoV-OC43, which both had lower seropositivities among HWs, do not follow this trend and have slightly higher seroprevalences in the uninsured HWs compared to the insured HWs.

Vaccines are available for two of the virus species with higher seroprevalence in our HW population (HAV and EV-C, which includes poliovirus), and therefore the differences we observed could be due to differences in rates of either natural infection or vaccination. Therefore, we further dissected the antibody responses against these viruses by examining protein– and peptide-level reactivity profiles. Notably, all HAV vaccines approved for use in the US are inactivated and it has been shown that antibody responses to natural infection and vaccination can be differentiated by measuring the response to nonstructural proteins, to which a response will only be generated with a natural infection (i.e., when there is virus replication and production of non-structural proteins) (42). Therefore, to examine the role of natural exposure to HAV in the observed difference in seropositivity, we mapped all enriched HAV peptides for each seropositive sample across the HAV proteome. We did not observe a significant difference between HW and NHW cohorts in the proportion of seropositive individuals with ≥1 enriched peptide from an HAV non-structural protein (Fisher’s exact test p-value=0.77). In fact, we observed high rates of reactivity against non-structural HAV proteins across both groups (HW=53/71, NHW=17/21) (Figure 5A). These findings indicate that most of the seropositive individuals in our study have likely been naturally infected by HAV and that the observed difference in seroprevalence between HW and NHW is probably not driven by differences in vaccination rate.

**Figure 5.**
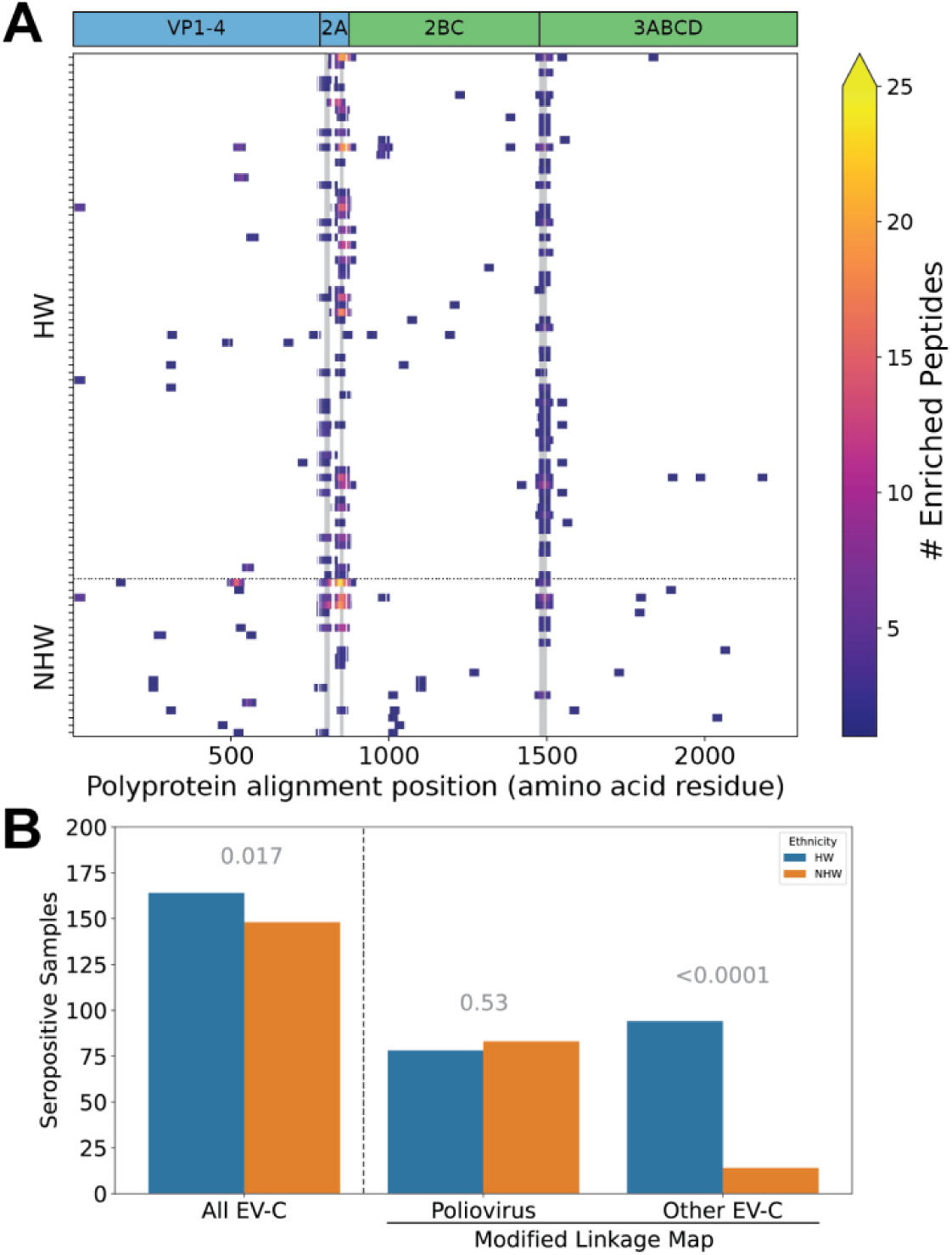
Ethnicity-based disparities in viral infection rates are not driven by vaccination against hepatitis A virus (HAV) or poliovirus. A) Enriched peptides and public epitopes identified in both structural and non-structural HAV proteins. Heatmap of enriched peptides across the HAV proteome for all samples seropositive for HAV with a Z score threshold of 15. Samples are broken up by ethnicity on the y-axis and position within the HAV polyprotein is shown on the x-axis (amino acid residues, alignment coordinates). The three most commonly reactive epitopes within these samples are highlighted with gray vertical markings. The positions of individual viral proteins are indicated across the top; blue=structural (vaccination or infection) and green=non-structural (infection only). B) Bar plot showing the number of seropositive HW (total n=198) and NHW (total n=198) individuals. Seropositivity was calculated using the original linkage map (Focal93) with poliovirus peptides assigned to the Enterovirus C (EV-C) virus species (“All EV-C”), or with a modified linkage map where peptides sharing ≥1 7mer with polioviruses were removed from the EV-C species and treated as their own category (“Poliovirus” and “Other EV-C”). P-values for binomial GLM comparing the impact of ethnicity (HW, NHW) on seropositivity are indicated in gray above each viral category.

Next, we sought to determine the role of poliovirus vaccination on the increased seropositivity to EV-C in HWs compared to NHWs. In this case, both inactivated and attenuated (replication competent) vaccines were used in the US prior to 2000, and both are still administered in Mexico, which borders Arizona and is the most common source of immigrants in the state (43). Therefore, instead of comparing protein-level patterns of antibody reactivity, we compared antibody reactivity to peptides designed specifically from the three strains of poliovirus. Specifically, we reran our estimates of seropositivity after separating the three polioviruses (UniProt Accessions can be found in Table S3) from the rest of EV-C. In this new analysis, we saw no significant difference in estimated serostatus for poliovirus (p-value = 0.53) with 78 (39%) HW and 83 (42%) NHW positive samples (Figure 5B). However, we did observe a highly significant difference in seropositivity between HWs and NHWs for “Other EV-C” strains, with 94 (47%) and 14 (7%) seropositive samples, respectively (p-value<0.0001) (Figure 5B). These results indicate that the observed disparity in EV-C seropositivity between HWs and NHWs is not driven by differences in natural infection or vaccination with poliovirus but is likely caused by differences in infection rate with other EV-C viruses.

To determine whether there were particular non-polio EV-C viruses that were driving the disparity seen in the “Other EV-C” group, we further analyzed the reactive peptides assigned to this group. First, we split the 27 International Committee on Taxonomy of Viruses (ICTV) listed EV-C isolates into six phylogenetic clades (Figure 6A, one clade includes only the polioviruses). Next, we assigned each “Other EV-C” peptide to the most similar EV-C clade (based on shared amino acid 7mers) and assigned these peptides scores equivalent to the number of contained 7mers that are unique to that clade. We then calculated relative peptide scores for each EV-C clade by summing the scores for 1) all “Other EV-C” peptides in the full HV1 PepSeq library (null distribution) and 2) the subsets of enriched “Other EV-C” peptides observed in the HW and NHW samples that were seropositive for “Other EV-C” (Figure 6B, Supplemental Figure 3). We observed antibody reactivity to peptides assigned to all five “Other EV-C” clades, and the clade-specific relative peptide scores varied substantially between individuals (Supplemental Figure 3). These results suggest that a variety of different EV-C viruses may be contributing to the observed disparity in EV-C seropositivity between HWs and NHWs. However, some of these clades (e.g., EV-C_2 and EV-C_3) may be more common than others, based on differences in the relative peptide scores between the expected (“Full Library”) and observed (“NHW”, “HW”) distributions (Figure 6B).

**Figure 6.**
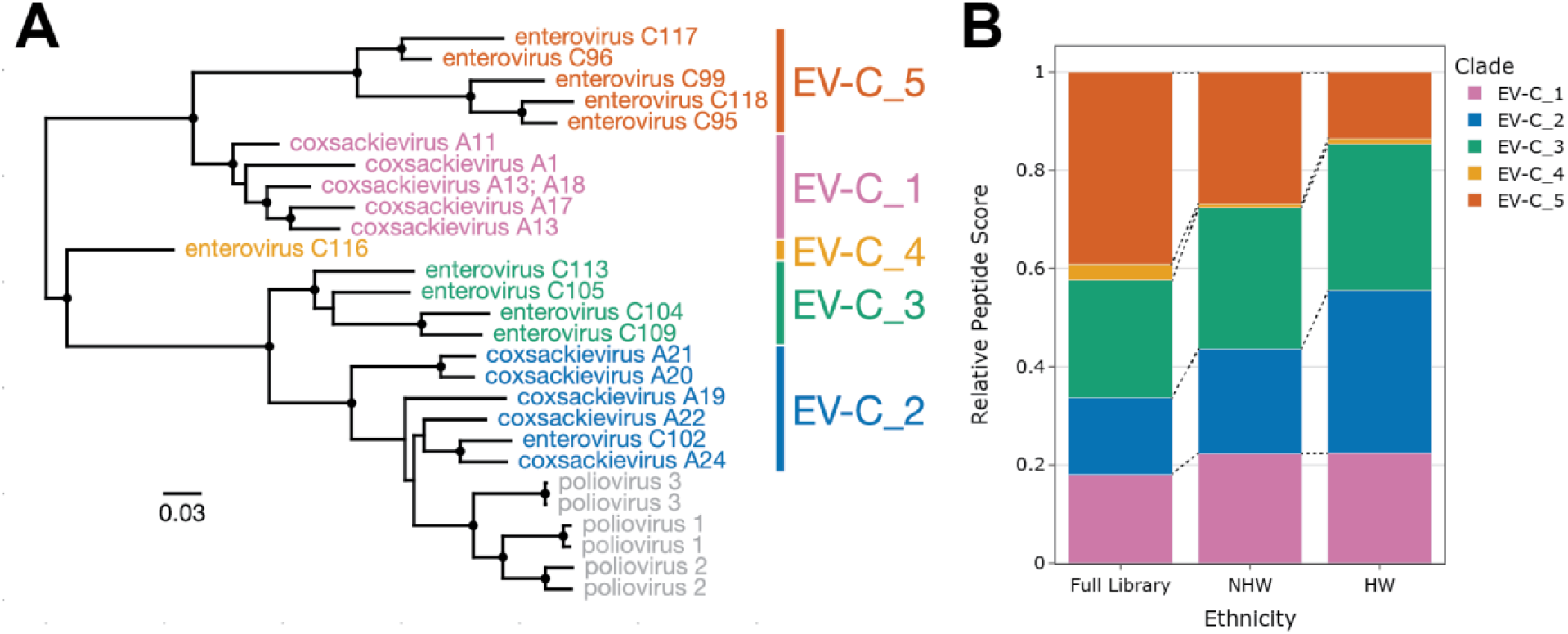
Antibody reactivity against clade-specific EV-C peptides. A) Maximum-likelihood phylogenetic tree of all 27 ICTV listed EV-C isolates based on an amino acid alignment of the full polyprotein. Branch lengths indicate relative levels of amino acid divergence with the scale bar indicating the equivalent of 0.03 changes per site. Black circles indicate nodes with bootstrap support ≥80. Six phylogenetic clades are identified by the different colors. Genbank accession numbers for each sequence can be found in Table S7. B) Bar plot showing clade-specific relative peptide scores for all “Other EV-C” peptides present in the HV1 PepSeq library (Full Library, null distribution) compared to the subset of enriched peptides for all HW and NHW individuals predicted to be seropositive against “Other EV-C” (Figure 5B). Each color represents one of the “Other EV-C” phylogenetic clades shown in (A).

## Discussion

In this study, we used PepSeq, a highly-multiplexed serology platform, to broadly assess virus infection histories and identify differences in seropositivity among various subsets of the population served by Valleywise Health in Phoenix, AZ. PepSeq allows for 100,000s of peptide antigens to be simultaneously assayed for antibody reactivity, and thus, it has the potential to facilitate the comprehensive characterization of differences in viral infection rates among subsets of a community. In contrast, the singleplex nature of traditional serological techniques (e.g., ELISA) has required that previous studies focus on a small number of high priority viruses (44, 45).

In general, our results were consistent with expectations from previously published studies. First, we observed a general trend toward higher seropositivity with increasing age, a pattern that has been reported for a variety of viruses (32–34, 39). In fact, we observed this pattern at two levels: 1) a positive correlation between an individual’s age and the number of seropositive virus species called (Figure 3B) and 2) negative age-associated GLM coefficients for most viruses, indicative of higher seroprevalence with increased age (Supplemental Figure 2). Second, we found that overall estimates of seroprevalence for individual viruses were broadly consistent with expectations from molecular and singleplex serological surveys. For example, with a Z score threshold of 15, seven viruses had estimated seropositivities ≥90% (Figure 3C). Among these were five common respiratory viruses, for which the expected seroprevalence is near 100% by adulthood (46): human rhinoviruses A, B and C, human orthopneumovirus (aka respiratory syncytial virus) (47) and human respirovirus 3 (aka human parainfluenza virus 3) (48). Also included in this set are human gammaherpesvirus 4 (aka Epstain-Barr virus) and Norwalk virus (aka norovirus), consistent with published serological surveys of adults in the US (33, 38). We also found that relative seroprevalence estimates for closely related viruses were generally consistent with documented differences in abundance. For example, while we estimated a seroprevalence for the more common human respirovirus 3 of 91%, the less common “parainfluenza” viruses (human respirovirus 1 and rubulaviruses 2 and 4) were estimated to have seroprevalences of 25-36% (49). Similarly, we estimated the seroprevalence of HSV-1 (85%) to be ∼2.2x higher than that of HSV-2 (38%) (Figure 3D), reflecting known differences in prevalence of these related viruses (50). Taken together, these observations provide strong support for the use of our highly-multiplexed serology approach for broadly characterizing virus infection histories.

Although we did not observe any statistically significant differences in seropositivity between males and females in our study, we did observe significant differences between HWs and NHWs for seven different viruses, and these differences were largely consistent across different Z score thresholds for peptide enrichment (Figure 4A). For five of these viruses (CMV, HSV-1, HAV, SaV-A and HAdV-D), we observed significantly higher seropositivity within our HW subpopulation, while the other two (HHV-7 and PeV-A) showed higher seropositivity in our NHW subpopulation, and several of these significant trends are consistent with published studies, lending additional credibility to our highly-multiplexed approach. For example, we detected higher seropositivity among HWs for both CMV (10-20% higher than among NHWs) and HSV-1 (10-30% higher), consistent with the results of previous US population studies that used singleplex antibody assays (32, 50). We also observed significantly higher seropositivity for HAV among HWs, which is consistent with published rates of seropositivity in the US, as well as known differences in seropositivity between the US and Mexico (34, 51–53).

We also observed several previously undocumented disparities in infection history between our HW and NHW populations, most of which involved viruses that are rarely targeted in serosurveys. One example is the recently described picornavirus SaV-A, which had ∼10% higher seropositivity within our HW subpopulation. Little is known about the seroprevalence of SaV-A in the US, however, it has been documented in wastewater samples from multiple US states, and a study in Arizona found evidence of SaV-A in 15% of wastewater samples (54, 55). Another previously undocumented disparity involves HAdV-D, for which we observed 20-30% higher seropositivity within our HW subpopulation (Figure 4). HAdV-D is a highly diverse species that has been shown to cause severe disease such as epidemic keratoconjunctivitis in immunocompromised individuals. However, little is known about the general health impact of HAdV-D, and therefore, it is difficult to deduce the impact this virus might have on the wellbeing of this population (56). Finally, we estimated significantly higher seroprevalence in our NHW subpopulation for two understudied viruses: PeV-A and HHV-7. PeV-A is a widespread virus that is associated with respiratory and gastrointestinal symptoms (57–59). However, certain strains of PeV-A are associated with more severe disease, such as meningitis and sepsis-like illness in infants (58). HHV-7 is a highly prevalent virus, which is primarily contracted in early childhood, resulting in life-long latent infections that typically remain asymptomatic. However, HHV-7 has also been linked with febrile seizures and as a possible cause of encephalitis (60–62). More work is needed to understand the full impact that these undocumented disparities may have on human health within these populations.

Arizona is home to a large and diverse immigrant population and this has likely contributed to our observed differences in seropositivity among ethnicities. According to estimates from 2018, immigrants (i.e., foreign-born individuals) comprised 13% of the population in Arizona, 16% of the state’s population were native-born Americans with at least one immigrant parent, and 55% of all immigrants in Arizona were from Mexico, with the next most common countries of origin (Canada, India, Philippines) each accounting for only 4% of the immigrant population (63). Although we did not have access to the immigration status for the individuals included in this study, we were able to examine the payor source associated with each individual’s medical visit, and we used a lack of insurance coverage (i.e., “self pay”) as a proxy for immigration status. In other words, it was assumed that the uninsured population would contain a higher proportion of immigrants compared to the insured population, and consistent with this assumption, we observed a much higher proportion of uninsured individuals within our HW subpopulation compared to our NHW subpopulation (Figure 1B).

By comparing seropositivity estimates between the uninsured and insured HW subpopulations, we were able to demonstrate that, for the majority of viruses that exhibited significant or near significant differences in seropositivity between ethnicities, the insured HW population represented an intermediate level of seropositivity between NHWs and uninsured HWs (Figure 4B). This pattern is consistent with the hypothesis that a substantial portion of the differences in seroprevalence between ethnicities can be attributed to differences in seroprevalence between immigrant and non-immigrant populations. Antibody responses to viruses can be long-lived. Therefore, one possible explanation is that we are seeing evidence of differences in virus exposure rates, possibly during adolescence, for those who were raised in the US versus those who were raised outside of the US, potentially in less developed, resource-limited countries. Many of the viruses flagged by our analysis are commonly encountered during childhood, and several are known to be more prevalent outside of the US (CMV, HSV-1, and HAV) (64–66). However, other factors, such as differences in the dynamics of virus transmission in immigrant communities within the US could also be contributing to the observed patterns.

Virus transmission mechanisms may also play an important role in determining which viruses are most likely to be associated with disparities among ethnic groups. Among the seven viruses with a significant difference in seroprevalence between HWs and NHWs, three are primarily transmitted through intimate contact (CMV, HHV-7, HSV-1), three through the fecal-oral route (HAV, PeV-A, SaV-A) and one (HAdV-D) has been associated with a variety of transmission routes, including close personal contact, the fecal-oral route, and respiratory droplets. Notably absent from this list is respiratory viruses that are primarily spread through airborne transmission, which make up roughly 23% of the focal93 viruses (Table S4). This absence of airborne transmitted viruses is likely due to the wider potential radius for spread from person to person and suggests that viruses that rely on close contact for successful transmission are more likely to be associated with population-level disparities.

Among our differentially seropositive species, two (HAV and EV-C) include viruses for which there are widely available vaccines, and for both of these, our highly-multiplexed assay allowed us to assess the relative roles of vaccination and natural infection on the observed differences in seropositivity. For HAV, we were able to differentiate antibody reactivity from vaccination and natural infection by leveraging the ability of PepSeq to simultaneously measure antibody reactivity across multiple HAV protein targets. Specifically, we compared patterns of antibody reactivity between the structural and non-structural proteins of HAV (Figure 5A). For nonstructural proteins to be produced, active replication of viral particles must occur (67). However, all available vaccines in the US contain inactivated viruses, and therefore, vaccination will not elicit antibodies that target nonstructural proteins. Our results show high levels of antibody reactivity against nonstructural HAV proteins in both HW and NHW individuals. This suggests that the higher seroprevalence among HWs is not because of higher vaccination rates in this population.

There is also a commonly used vaccine that includes three viruses that belong to the EV-C species — poliovirus 1, 2, and 3 — and both inactivated and live attenuated versions were in use during the lifetimes of our participants. Therefore, to control for differences in antibody responses that may be driven by differences in rates of vaccination, we separated our EV-C peptides into two categories: those that share at least one 7mer with any of the three polioviruses (i.e., those most similar to vaccine antigens) and those that don’t. We saw no difference between our ethnicities in reactivity against the peptides most likely to be recognized by vaccine-induced antibodies, but a highly significant difference in reactivity against “Other EV-C” peptides (40% higher seropositivity among HWs; Figure 5B). Further analysis showed that the observed antibody reactivity profiles are consistent with past exposures to a wide variety of EV-C viruses (Supplemental Figure 3), but that some phylogenetic clades may be contributing more than others to the observed disparity in EV-C infections between HWs and NHWs (Figure 6B). Notably, while EV-C_5 peptides are overrepresented in our starting library, they are comparably underrepresented among our enriched peptides, especially in our HW subpopulation (Figure 6B). Interestingly, EV-C_5 has been predominantly isolated from nasal/throat swabs and nasopharyngeal aspirates, suggesting these viruses are likely transmitted via respiratory droplets (68, 69). In contrast, the other EV-C clades have been isolated almost universally from stool samples, indicating that they primarily infect the gastrointestinal tract and are likely to be spread through the fecal-oral transmission pathway (68, 69). This tissue specific tropism aligns with our general hypothesis that the population level virus infection disparities are more commonly driven by viruses that require close contact transmission routes.

## Conclusion

Overall, our highly-multiplexed serology assay was successful in broadly characterizing antiviral antibody reactivities and allowed us to infer individual infection histories across the virome. The recapitulation of several known differences in seropositivity between two ethnic groups provides confidence in the quality of our analysis and highlights promising future applications for this type of approach. Additionally, our study revealed several previously undocumented disparities in virus seropositivity between HWs and NHWs in our study population. Future studies will be needed to better understand the clinical significance of these differences in infection rates, and to develop medical and social interventions to minimize the impact of these disparities. Our results also demonstrate the potential for highly-multiplexed serology to finely dissect the specificity and breadth of antibody responses, thus enabling an unprecedented view into an individual’s history of infection.

## Materials and Methods

### Study population and sample collection

In total, 400 serum samples were obtained from Valleywise Health in Phoenix, AZ. These samples were collected in late May and early June 2020. They were remnant samples initially collected as a part of the patients’ standard of care, and were collected from several different facilities and in a variety of contexts including outpatient encounters (55.5%), inpatient encounters (35%) and emergency department visits (9.5%). Researchers at Northern Arizona University (NAU) did not have access to any identifiable patient information. This study was reviewed and approved by the Valleywise Health and NAU Institutional Review Boards (approval number 1545420).

To maximize statistical power to detect differences in seroprevalence, our cohort for this study was equally divided among four sub-populations: HW males (n=100), HW females (n=100), NHW males (n=100) and NHW females (n=100). To minimize the effect of age in detecting differences in seroprevalence, we selected only individuals within an age range of 30-60 years old. Self-reported ethnicity, gender and age were the only characteristics considered for inclusion in this study. However, because Valleywise Health is a safety net hospital, we do not expect our study population to represent a random sampling of the population of Phoenix, AZ. Rather, it is likely to include a higher proportion of individuals with low income and from under-served populations. There is also some potential for bias associated with the use of remnant samples collected from patients actively receiving medical care (compared to a random sample of adults). However, given the wide variety of encounter types represented in our sample, we expect any impact to be minimal.

In total, 83% of the population served by Valleywise Health consists of racial and ethnic minorities, and at the Valleywise Health ambulatory clinics, 59% of patients are Hispanic. The majority of families served by Valleywise Health are at or below 150% of the Federal Poverty Level, and approximately 55% of Valleywise Health patients are enrolled in a government health insurance program for low income people, or have insufficient private insurance or no insurance.

For each patient, we also obtained payor source, as this can serve as a useful, though incomplete, indicator of socio-economic status. As the exact payor source is highly variable among individuals, we reduced the complexity of this categorical variable by assigning every individual to one of six general categories: 1) “Commercial”, which included all commercial health plans; 2) “Medicaid”, which included both Arizona Health Care Cost Containment plans and out-of-state Medicaid; 3) “Medicare”; 4) “Dual-SNP”, which included any dual special needs plans for individuals who qualify for both Medicaid and Medicare; 5) “Self Pay”, for individuals without insurance and 6) “Other”, which served as a final catch-all category that included funding through charitable organizations like the Ryan White HIV/AIDS Program and other government plans such as Tricare (Table S5).

### PepSeq Library Design and Assay

To broadly assess antiviral antibody reactivity, we utilized the PepSeq platform to perform highly-multiplexed peptide-based serology (29). Specifically, we used the human virome version 1 (HV1) library described in (26). In brief, the HV1 library consists of 244,000 unique DNA-peptide conjugates (i.e., PepSeq probes). The variable peptide portion of each molecule is 30 amino acids long and the peptides were designed to broadly cover potential linear epitopes present in the proteins of viruses known to infect humans. Libraries of these PepSeq probes are created through a series of bulk, *in vitro* enzymatic reactions (29).

Each assay was conducted as described in Ladner et al. (26). Broadly, the PepSeq assay involves the incubation of serum with a diverse pool of PepSeq probes. Immunoglobulin G (IgG) is then precipitated using magnetic protein G beads, non-binding PepSeq probes are washed away, and the relative abundance of each probe is quantified using PCR and high-throughput sequencing of the DNA portion of the molecules (29). Specifically, 5μL of a 1:10 dilution of serum in Superblock T20 (Thermo) was added to 0.1 pmol of the PepSeq library for a total volume of 10 uL and was incubated at 20°C overnight. The binding reaction was incubated with pre-washed protein G-coated beads (Thermo) for 15 minutes, after which the beads were hand washed 11 times with 1x PBST. After the final wash, beads were resuspended in 30μL of water and heated to 95°C for 5 minutes to elute the bound PepSeq probes. Elutions were amplified and indexed using barcoded DNA oligonucleotides (Table S6). Following PCR, a standard bead cleanup was performed and products were individually quantified (Quant-It, Thermo Fisher), pooled, re-quantified (KAPA Library Quantification Kit, Roche) and sequenced on a NextSeq instrument (Illumina). For this study, each sample was assayed in duplicate and ≥1 buffer-only negative controls were included on each 96-well assay plate. Potential batch effects were controlled through equal representation of each of our four focal subpopulations on each plate, as well as through the inclusion of negative controls from all plates in read count normalization and the generation of the peptide bins.

### PepSeq Analysis

We used PepSIRF v1.5.0 (70, 71) to analyze the high-throughput sequencing data. Demultiplexing and assignment of reads to peptides was done using the *demux* module of PepSIRF allowing up to 1 mismatch within each of the index sequences (12 and 8 nt, respectively) and up to 3 mismatches with the expected DNA tag (90 nt). Z scores were calculated using the *zscore* module of PepSIRF, which implements a method adapted from (72). This process involved the generation of peptide bins, each of which contained ≥300 peptides with similar expected abundances in our PepSeq library. Expected abundance for each peptide was estimated using buffer-only negative controls. In total, 12 independent buffer-only controls from 7 different assay plates were used to generate the bins for this study. The raw read counts from each of these controls were first normalized to reads per million (RPM) using the column sum normalization method in the *norm* module of PepSIRF. This served to normalize for differences in total sequencing depth between samples. Bins were then generated using the *bin* PepSIRF module. RPM counts for each peptide were then further normalized by subtracting the average RPM count observed within our buffer-only controls. This second normalization step was used to control for any differences in initial relative abundance among peptides contained within the same bin. Each Z score was calculated using peptides contained within the same bin and corresponds to the number of standard deviations away from the mean, with the mean and standard deviation calculated using the 95% highest density interval to exclude any enriched peptides.

The *enrich* module of PepSIRF was used to determine which peptides had been enriched through our assay (i.e., were bound by serum IgG isotype antibodies). This module identifies peptides that meet or exceed minimum Z score thresholds, in both replicates for each sample. Z score thresholds were selected to minimize the number of false positive calls of peptide enrichment (determined through the analysis of negative controls that were not considered in the formation of bins), and multiple Z score thresholds were examined to determine the sensitivity of our results to changes in this threshold.

The lists of enriched peptides were converted into lists of putative species-level seropositivities using the *deconv* module of PepSIRF. The goal of this module is to predict the minimum list of viruses to which an individual has likely been infected, while considering shared sequence diversity among different viruses. To accomplish this, the *link* module was first used to generate a linkage map that relates individual peptides to virus species. A link between a peptide and virus indicates that enrichment of the peptide could be explained by exposure to the linked virus species, and these links were made whenever a peptide shared ≥1 amino acid 7mer with a target protein sequence obtained from a particular virus species. Because of shared sequence diversity, a single peptide can be linked to multiple species, not just the species from which the peptide was designed. Furthermore, the strength of the link between peptides and viruses was quantified with scores that correspond to the number of shared 7mers. Therefore, the maximum link score was 24 and the minimum link score was 1.

The HV1 PepSeq library includes peptides derived from 390 virus species, but many of these viruses have only very rarely been associated with human infections (e.g., foot and mouth disease virus, simian foamy virus) and/or occur in distant and spatially restricted parts of the world (e.g., Ebola virus, Crimean-Congo hemorrhagic fever virus), and therefore it is very unlikely that our focal population would have been exposed to these viruses. To determine whether the inclusion of these viruses was impacting the results of our analysis, we generated two distinct linkage maps. The first (“Full”, Table S1) included all 390 viruses included as targets in the HV1 library design (26), while the second only included 93 virus species (“Focal93”, Table S1), excluding any viruses to which exposure within the study population was deemed to be very unlikely.

The *deconv* module of PepSIRF was then used with each of these linkage maps to identify the most parsimonious set of virus species that can explain each set of enriched peptides. The results, therefore, can be interpreted as potential seropositivities for each sample. The *deconv* module accomplishes this through an iterative process. In each round, species-specific scores are generated by summing the species-level scores from each enriched peptide, and the species with the highest score is selected for inclusion in the output. We set a score threshold of 40; a species must reach or exceed this score in order to be included in the output. Additionally, to account for related species with similar scores, we allowed for ties between species if the lower scoring species had a score ≥80% of the higher scoring species (--score_tie_threshold 0.8) and if ≥70% of the enriched peptides contributing to these scores were identical between species (–– score_overlap_threshold 0.7). In our analysis, we considered an individual to be seropositive for all tied species. Ties accounted for less than 2% of seropositivity calls. A separate *deconv* analysis was run for each Z score threshold.

### Identification of disparities

To identify significant differences in estimated seropositivity between ethnicities and/or genders, we utilized a generalized linear model (GLM) implemented in Python using statsmodels v3.8.8 (73). Specifically, for each viral species, we fit a binomial GLM with a single dependent variable (seropositivity; 0 or 1 for each individual) and three independent variables [ethnicity (categorical), gender (categorical) and age (continuous)]. We utilized an alpha of 0.05 for determining significance, along with a Bonferroni correction for multiple tests (i.e., number of viruses). To reduce the total number of tests, we only examined viruses with estimated population-level seropositivities between 5 and 95%. These thresholds were chosen based on power simulations, which indicated that, with our sample size, it would be unlikely to detect differences in seropositivity <10% as statistically significant.

### Subspecies analysis of reactivity profiles

To assign enriched HAV peptides to individual proteins, we first aligned all 360 HAV amino acid sequences from which HV1 peptides were designed using mafft v7.490 (74) with the *G-INS-i method. Annotations from one of these sequences (Uniprot:Q9DWR1) were then translated into alignment coordinates for the purposes of visualization (Figure 5A) and for assigning peptides to proteins. A peptide was considered structural if ≥25 peptide amino acids (83%) were assigned to HAV proteins VP1-4 and/or 2A (blue in Figure 5A). A peptide was considered non-structural if ≥25 peptide amino acids were assigned to HAV proteins 2BC and/or 3ABCD (green in Figure 5A).

To determine if the disparity in EV-C seropositivity was driven by differences in vaccination rate, we created a new linkage map where all peptides containing at least one 7mer from any of the 2,653 poliovirus sequences (Table S3) used in the creation of the HV1 library were assigned to a new taxonomic category called “poliovirus”. All EV-C peptides that did not share a 7mer with poliovirus sequences were assigned to an “Other EV-C” category. Next, we reran the *deconv* module using this modified linkage map to estimate seropositivity for these two categories separately. With this analysis, it is possible for an individual sample to be found seropositive for 1) just one of these categories, 2) both categories (if there are multiple enriched peptides from each category), or 3) neither of these categories (even for samples previously found to be seropositive for EV-C, if the enriched peptides are split between the new categories). This is a conservative approach for assessing the impact of poliovirus vaccination/infection because it assumes that any antibody recognizing a peptide that shares at least one amino acid 7mer with poliovirus was generated in response to poliovirus infection/vaccination. In reality, these responses could have been stimulated by many different EV-C strains.

To examine the contribution of different strains of EV-C to our observed “Other EV-C” seropositivities, we assigned each “Other EV-C” peptide to a single subspecies group based on shared amino acid 7mers, with each peptide assigned a score (between 1 and 24) equivalent to the number of contained 7mers that were unique to the assigned group. To improve the sensitivity of our analysis (i.e., the number of informative peptides), we focused on clades of related EV-C isolates. As of December 8, 2023, the ICTV website for the Enterovirus genus (https://ictv.global/report/chapter/picornaviridae/picornaviridae/enterovirus) listed NCBI GenBank accession numbers for 27 EV-C isolates, including 6 poliovirus isolates (Table S7). We downloaded polyprotein amino acid sequences for each of these and aligned them using mafft v7.490 (74) with default settings. We generated a maximum-likelihood phylogeny from this alignment using raxml-ng v0.5.1b (75) with the LG+FC+I+G8m model, which was selected as optimal using ModelGenerator v0.85 (AIC1) (76). Based on this phylogeny, we divided the EV-C subspecies into six phylogenetic clades, including five composed of non-poliovirus EV-C (EV-C_1-5) (Figure 6). For each enriched “Other EV-C” peptide, we determined the number of amino acid 7mers shared with the ICTV reference sequences from each EV-C clade. We assigned the peptide to the clade with the highest score and normalized the associated score for this peptide by subtracting the next highest clade-specific score (no assignment was made if two clades had identical 7mer scores). To calculate relative reactivity scores against the five non-poliovirus clades (“Relative Peptide Score” in Figure 6B, Supplemental Figure 3), each clade-specific sum of enriched peptide scores was divided by the total sum of scores across all clade-assigned peptides for that sample. For the ethnicity-level composite scores shown in Figure 6B (“HW” and “NHW”), we summed enriched peptide scores across all HW or NHW individuals, respectively, who were seropositive for our “Other EV-C” category. For these composites, individual peptide scores were counted once for every sample that exhibited enrichment (i.e., a single peptide could be counted multiple times if that peptide was recognized by antibodies in multiple samples).

## Acknowledgements

We would like to acknowledge Sarah Namdarian for help collecting the clinical samples used in this study. This work was supported by the National Institute on Minority Health and Health Disparities of the National Institutes of Health under Award Number U54MD012388 and the State of Arizona Technology and Research Initiative Fund (TRIF, administered by the Arizona Board of Regents, through Northern Arizona University). The content is solely the responsibility of the authors and does not necessarily represent the official views of the National Institutes of Health.

## Data Availability

The raw peptide counts, linkage maps and data related to the figures from this study have been deposited in the Open Science Framework (https://osf.io/gvpzu/), DOI: 10.17605/OSF.IO/GVPZU. All custom code is available via GitHub (https://github.com/LadnerLab). Any additional information required to reanalyze the data reported in this paper is available upon request.

## Notes

### Competing Interest Statement

The authors have declared no competing interest.

### Author Declarations

This study was reviewed and approved by the Valleywise Health and NAU Institutional Review Boards (approval number 1545420).

